# A description of Lineage 1 *Mycobacterium tuberculosis* from Papua, Indonesia

**DOI:** 10.1101/2024.05.07.24307031

**Authors:** Hanif A. K. Djunaedy, Clarissa A. Febinia, Raph L. Hamers, Kevin Baird, Iqbal Elyazar, Nguyen Thuy Thuong Thuong, Hidayat Trimarsanto, Safarina G. Malik, Guy Thwaites, Reinout van Crevel, Bachti Alisjahbana, Lidya Chaidir, Philip M. Ashton

## Abstract

Indonesia has the third highest number of tuberculosis (TB) patients infected with Mycobacterium tuberculosis (*MTB*) Lineage 1 (L1). Most of these MTB L1 cases can be found in Indonesia’s remote easternmost province of Papua, one of Indonesia’s most underdeveloped provinces with a particularly high burden for TB. In this study, we sequenced and described 42 *MTB* L1 isolates from a well-characterized cohort of patients. We found a genetically diverse *MTB* L1 population with no association between pathogen genetic relatedness and place of residence or pathogen genetic relatedness and patient ethnicity, which could reflect mixing between different locales and ethnicities or our low sampling fraction. Only a small number showed genetic variants associated with drug resistance (5/42, 11.9%), probably due to a lack of effective treatment programs. The Papuan isolates showed similarities to other Island Southeast Asian Countries due to the high proportion of L1.2.1.2.1 (30/42, 71.4%), especially East Timor and the Philippines. This study fills a research gap of MTB L1 in Indonesian Papua and should serve as a stepping stone for further research in the region.

## Introduction

Tuberculosis remains a major global health concern, caused by bacteria in the *Mycobacterium tuberculosis* complex that can be categorized into at least eight lineages. While the modern *MTB* lineages, such as lineage 2 and lineage 4, are the most globally widespread [1], the deeper branching lineage 1 (L1), is mainly restricted to countries around the Indian Ocean [2]. Globally, around 28% of TB infections are caused by *MTB* L1 and 80% of these cases occur in India, the Philippines, Indonesia, and Bangladesh [3]. While the common ancestor to *MTB* is believed to have evolved in East Africa, *MTB* L1 is believed to have emerged following a migration to India between the 3rd and 4th millennium BCE and eventually spread to South Asia, Southeast Asia, and even returned to East Africa through human migration that was facilitated by the trans-Indian Ocean trade route [4].

Whole genome sequencing analysis has allowed for the categorization of *MTB* into sublineages [5]. *MTB* L1 has five major sublineages, L1.1.1, L1.1.2, L1.1.3, L1.2.1, and L1.2.2 [3]. A majority of L1.1.1 cases are from Mainland Southeast Asia, mainly Vietnam [2]. L1.1.2 can be found across Africa to Mainland Southeast Asia but is most widespread in India [2] while L1.1.3 is mainly restricted to the Indian subcontinent, particularly Bangladesh and Myanmar [3]. L1.2.2 can be found across East and Southeast Asia [2]. L1.2.1 is the most abundant *MTB* L1 sublineage, making up 30% of all *MTB* L1 cases in 2018 [3]. This sublineage can be found across Southeast Asia but has a particularly high presence in Island Southeast Asia, such as the Philippines and Indonesia [6].

Indonesia is classified by the WHO as a high-burden country for TB [7] and has the third highest burden for *MTB* L1 [3]. Most of the *MTB* L1 burden comes from Eastern Indonesia due to a higher incidence of *MTB* L1 in the east compared to the west [8–10]. Papua, Indonesia’s easternmost province, has a particularly high burden, caused by a lack of healthcare infrastructure, low levels of economic development, and high HIV prevalence [11,12]. Along with Papua’s remoteness and isolated communities, this has all contributed to the lack of knowledge regarding the genetic makeup and population structure of *MTB* in the province.

In this study, we attempt to fill the research gap regarding genomic understanding of L1 in Eastern Indonesia by describing a novel collection of *MTB* L1 from Indonesian Papua.

## Methods and Materials

### Study setting and sampling

Papua is Indonesia’s easternmost province, encompassing western New Guinea Island and bordering Papua New Guinea to the east. The provincial capital and largest city, Jayapura, sits on the northeastern corner of the province. The city is divided into five districts. Jayapura Utara, Jayapura Selatan, and Heram to the west are more urbanized compared to the semiurban Abepura to the south and Muara Tami to the east. Because of Jayapura’s greater economic development compared to the surrounding regencies, which is mostly rural, many come to the city for work or seek medical treatment and reside in the city temporarily.

Papua is demographically distinct from the rest of Indonesia. Unlike other Indonesian islands, where the population primarily descends from Austronesian peoples that arrived 3,000-4,000 years ago, indigenous Papuans first settled the island around 50,000 years ago [13]. *MTB* L1 was also found to be more likely associated with patients with ethnic backgrounds from Eastern Indonesia, mainly Maluku and Papua [14].

At the time of sampling between March 2011 and September 2012, Jayapura was served by two public hospitals and eleven community health centers. We recruited patients who have been consecutively diagnosed with smear-positive TB at both hospitals and three primary health centers. These isolates were collected as part of a previous study [14,15], which was approved by the ethics review committees at the National Institute of Health Research and Development, Indonesian Ministry of Health, Jakarta, Indonesia. Patients’ sociodemographic and clinical data were collected using a questionnaire and isolate lineages were determined through lineage specific markers in a bead-based multiplex ligation-dependent probe amplification (MLPA) assay [14,15]. Of the 66 L1 samples collected in this study, only 46 samples underwent successful reculturing, DNA isolation, and whole genome sequencing. Four isolates were determined to be non-L1 by TB-Profiler and did not undergo further analysis.

### DNA Extraction and Sequencing

DNA extraction was done at the Universitas Padjadjaran Faculty of Medicine in Bandung, West Java, Indonesia from culture using cetyl trimethylammonium bromide (CTAB) method, which followed a similar protocol to those described in Chaidir, *et al*., [15] and Chaidir, *et al*., [14]. Library preparation was done using a Nextera XT Kit and underwent whole-genome sequencing using an Illumina NextSeq system (Illumina, San Diego, CA, USA) at the Eijkman Institute for Molecular Biology in Jakarta, Indonesia.

### Bioinformatics and phylogenetic analysis

The raw reads were trimmed using trim-galore v0.6.7 [16], a trimming tool that implements Cutadapt and FastQC, and underwent analysis using the PHEnix SNP calling pipeline [17]. This pipeline uses BWA-MEM to map the trimmed reads to a reference genome [18] and uses the GATK function UnifiedGenotyper as a variant caller [19]. The *Mycobacterium tuberculosis* H37Rv genome (accession number NC_000962.3) was used as a reference. The reads were also analyzed using the TB-Profiler v4.3.0 tool to identify drug resistance mutations and sublineage information [20].

The resulting VCF files containing the SNPs from each isolate were converted into a consensus FASTA file representing the reference genome modified with the high-quality SNPs (>90% consensus, 5x depth, >30 minimum base quality), and missing positions (consensus < 90%, <5x depth or <30 minimum base quality) identified in each genome, also using the PHEnix tool. The consensus genomes for each isolate were combined into one FASTA file, and then underwent phylogenetic analysis. IQ-Tree v2.2.0.3 [21] was used for the analysis, with model selection by ModelFinder [22] which generates a consensus tree using the maximum-likelihood method and ultrafast bootstrapping approximation using UFBoot2 [23]. The resulting consensus tree was visualized and annotated using iTOL v6 [24]. The combined FASTA file was also used to calculate pairwise SNP distance using snp-dists v0.8.2 [25]. Other visualizations were performed using ggplot2 v3.4.2 [26] in R v4.3.1 [27].

### Comparison with Global Isolates

The Papua isolates were compared with a collection of global lineage 1 isolates that was obtained from the Sequence Read Archive and the European Nucleotide Archive. The run IDs and location of origin were obtained from several published works [2,28–30]. These isolates were pooled together and, when available, 20 isolates were chosen at random from each continent (**Table S1** and **Table S2**). For South and Southeast Asian countries, 20 isolates were chosen at random from each country when available. A total of 306 readsets were selected and underwent the same bioinformatics analysis as the Papua isolates. The lineage of each genome in NCBI SRA was downloaded from https://tbdr.lshtm.ac.uk/sra/browse and the proportion of the genomes from each country that were L1.2.1.2.1 was calculated.

## Results

Of the 66 samples from Papua that were inferred to be L1 based on previously generated data from MLPA assays [14,15], 42 were successfully sequenced and underwent further analysis. Based on the patient questionnaires [14,15], most of the patients sampled were male (29/42, 69.0%) and tended to be younger, with a median age of 28.5 and more than half (23/42, 54.8%) were under the age of 30 (**Table 1**). Thirty-two patients were residents of Jayapura City (76.2%). Of these, most resided in Jayapura Utara (15/42, 35.7%), with Jayapura Selatan in second (13/42, 30.9%), followed by Abepura (3/42, 7.1%), and only one patient resided in Muara Tami (2.4%). The rest (10/42, 23.8%) reside in other regencies across northern Papua, including Biak-Numfor (3/42, 7.1%), Yapen Islands (1/42, 2.4%), Nabire (1/42, 2.4%), Sarmi (1/42, 2.4%), and Jayapura Regency (4/42, 9.6%) (**Table 1** and **Figure 1**). In terms of ethnicity, Coastal Papuans made up the majority ethnic group sampled (29/42, 69.0%) while only two highland Papuans were sampled (4.8%). The rest (11/42, 26.2%) belong to ethnic groups originating from outside New Guinea Island, mostly from neighboring Sulawesi (9/42, 21.4%) (**Table 1**).

**Table 1:**
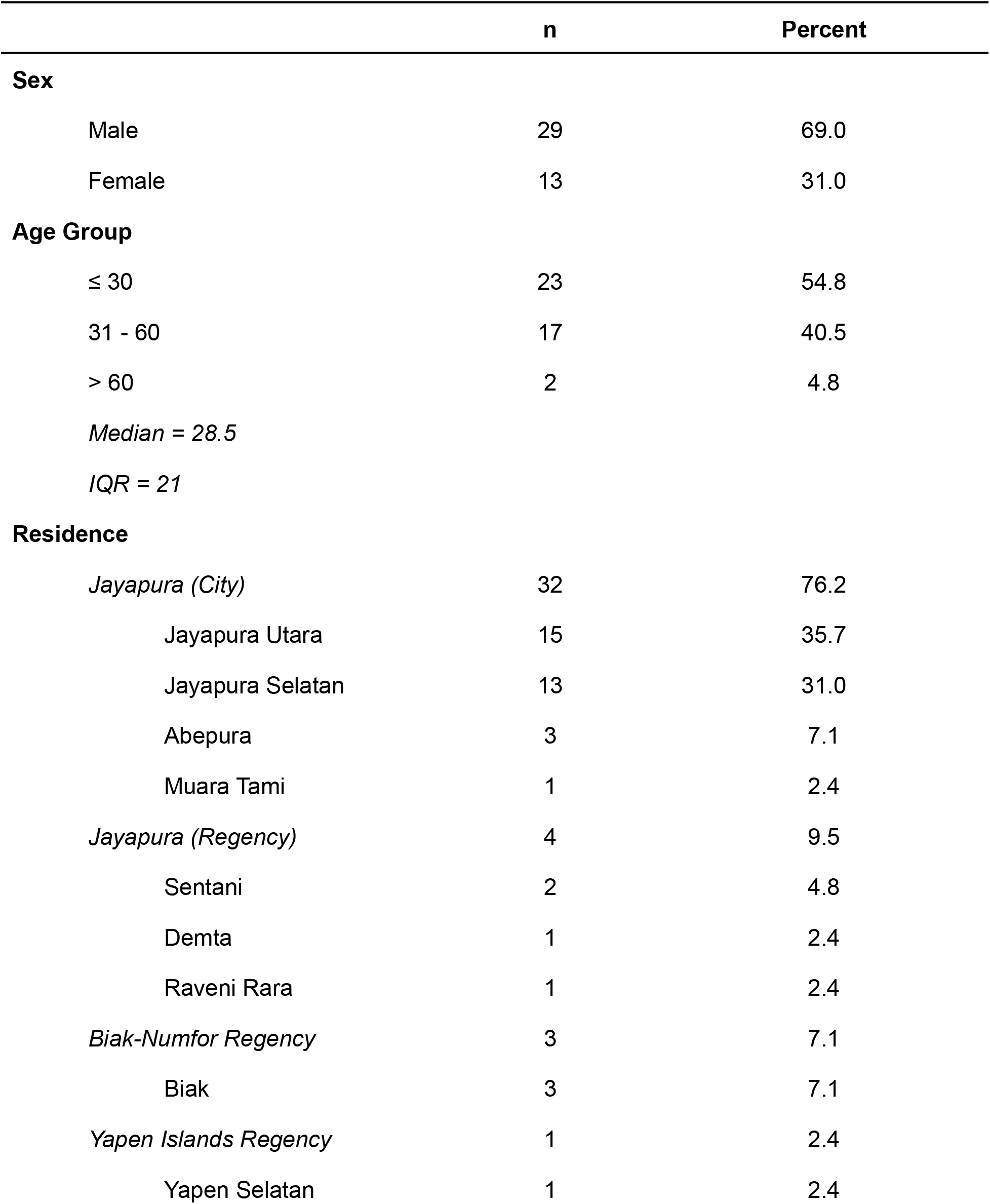
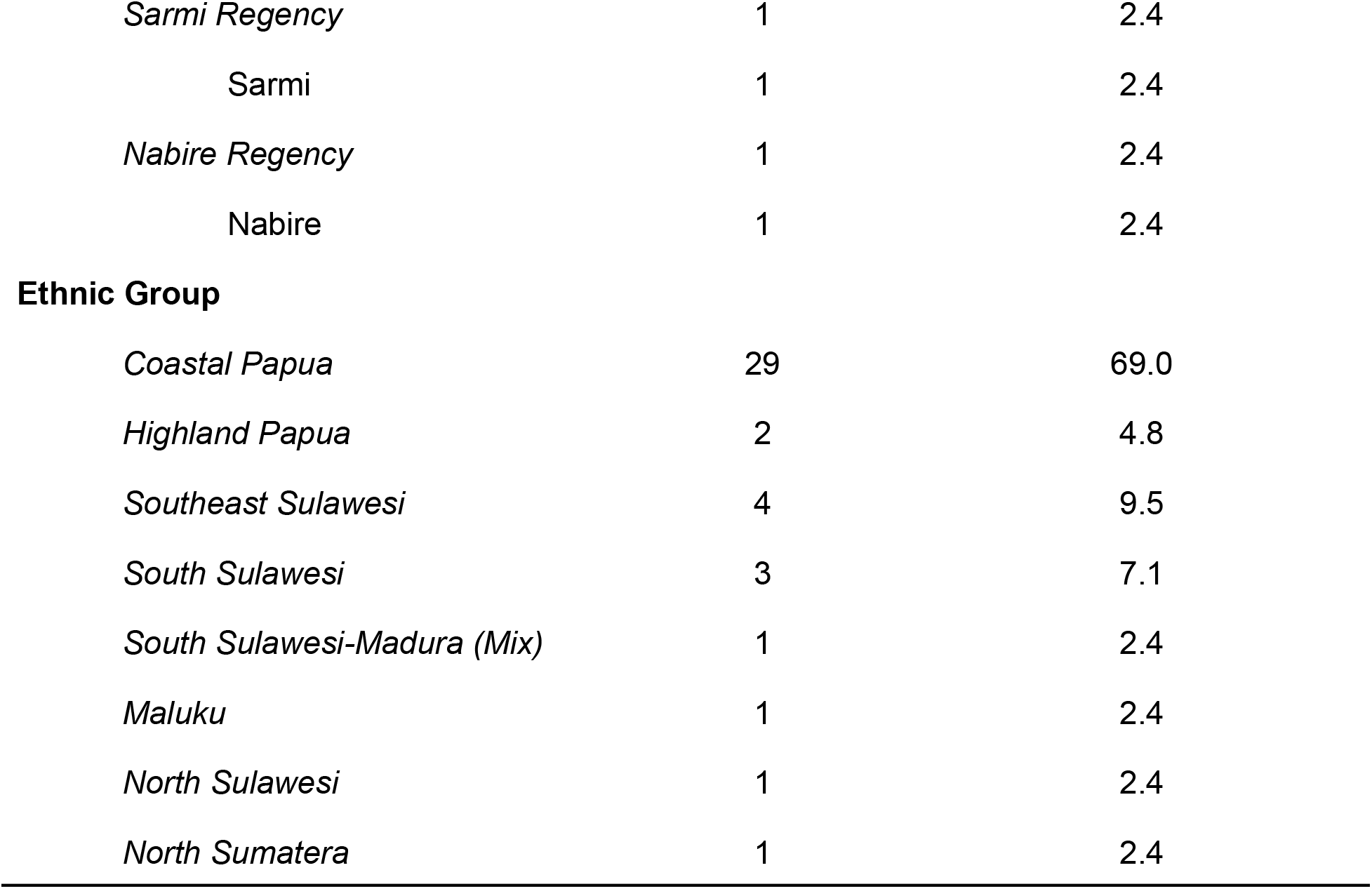
Patient Characteristics.

|  | n | Percent |
| --- | --- | --- |
| **Sex** |  |  |
| Male | 29 | 69.0 |
| Female | 13 | 31.0 |
| **Age Group** |  |  |
| ≤ 30 | 23 | 54.8 |
| 31 - 60 | 17 | 40.5 |
| > 60 | 2 | 4.8 |
| *Median = 28.5* |  |  |
| *IQR = 21* |  |  |
| **Residence** |  |  |
| *Jayapura (City)* | 32 | 76.2 |
| Jayapura Utara | 15 | 35.7 |
| Jayapura Selatan | 13 | 31.0 |
| Abepura | 3 | 7.1 |
| Muara Tami | 1 | 2.4 |
| *Jayapura (Regency)* | 4 | 9.5 |
| Sentani | 2 | 4.8 |
| Demta | 1 | 2.4 |
| Raveni Rara | 1 | 2.4 |
| *Biak-Numfor Regency* | 3 | 7.1 |
| Biak | 3 | 7.1 |
| *Yapen Islands Regency* | 1 | 2.4 |
| Yapen Selatan | 1 | 2.4 |
| *Sarmi Regency* | 1 | 2.4 |
| Sarmi | 1 | 2.4 |
| *Nabire Regency* | 1 | 2.4 |
| Nabire | 1 | 2.4 |
| **Ethnic Group** |  |  |
| *Coastal Papua* | 29 | 69.0 |
| *Highland Papua* | 2 | 4.8 |
| *Southeast Sulawesi* | 4 | 9.5 |
| *South Sulawesi* | 3 | 7.1 |
| *South Sulawesi-Madura (Mix)* | 1 | 2.4 |
| *Maluku* | 1 | 2.4 |
| *North Sulawesi* | 1 | 2.4 |
| *North Sumatera* | 1 | 2.4 |

**Figure 1:**
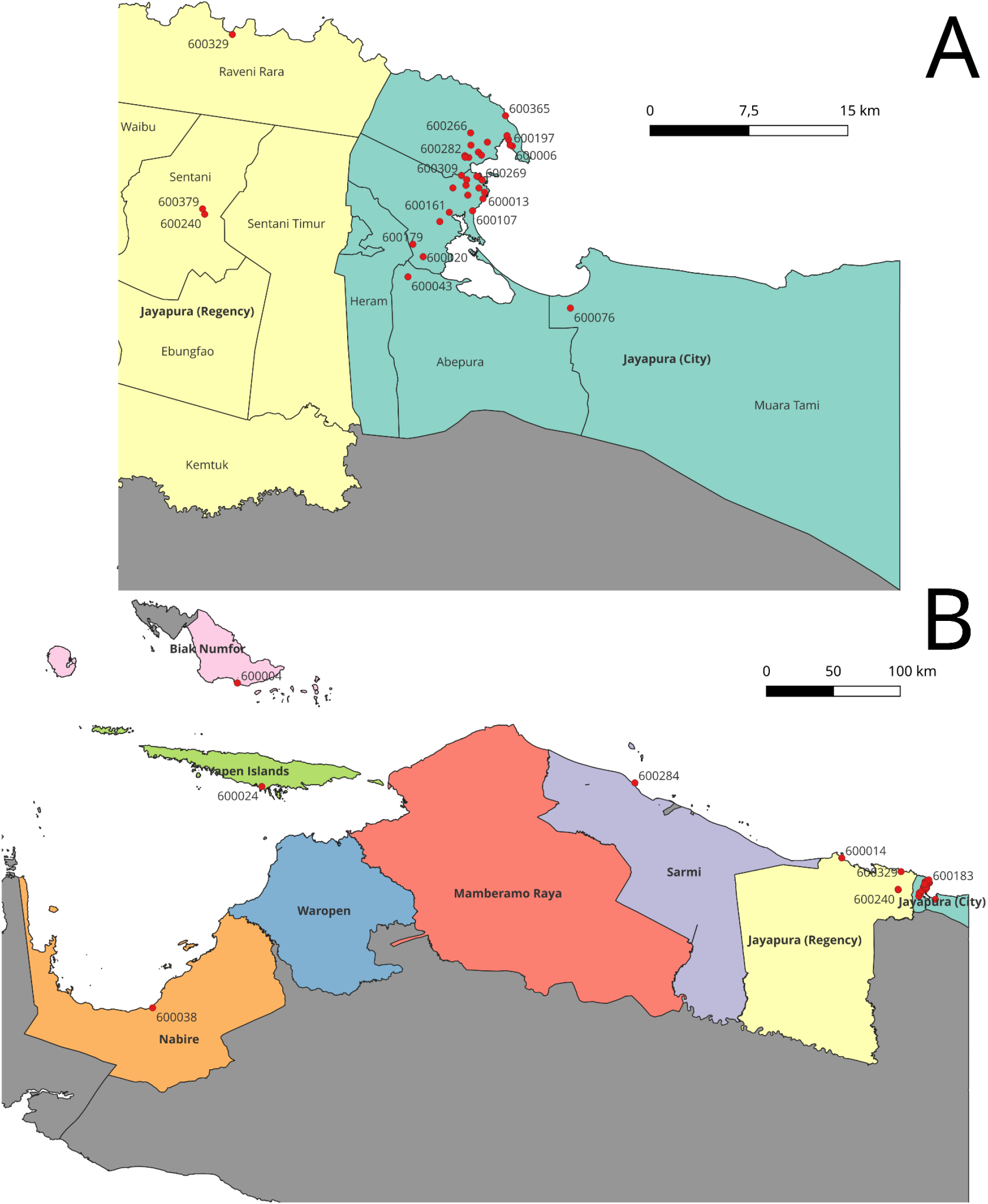
Distribution of patient residence in A) Jayapura City and Jayapura Regency and B) Northern Papua

A majority of the Papuan isolates belonged to L1.2.1.2.1 (30/42, 71.4%) (**Table 2**) and most of the Papuan isolates could be grouped with the clade based around sublineage L1.2.1 in the phylogenetic tree of global L1 isolates (**Figure 2**). Investigation of this lineage in *MTB* genomes in the Sequence Read Archive showed that the top 6 countries for proportion of L1.2.1.2.1 were; The Philippines (129/183, 71%), Malaysia (2/6, 33%), USA (26/220, 12%), Canada (195/1921, 11%), Lebanon (1/10, 10%), and Thailand (111/1507, 7%).

**Table 2:** Sublineage information of the Papua isolates. Sublineage prediction was generated through analysis by TB-Profiler.

| Sublineage | n | Percent |
| --- | --- | --- |
| Lineage 1 | 1 | 2.4 |
| Lineage 1.1.1 | 1 | 2.4 |
| Lineage 1.2.1 | 6 | 14.3 |
| Lineage 1.2.1.2 | 1 | 2.4 |
| Lineage 1.2.1.2.1 | 30 | 71.4 |
| Lineage 1.2.2.2 | 3 | 7.1 |

**Figure 2:**
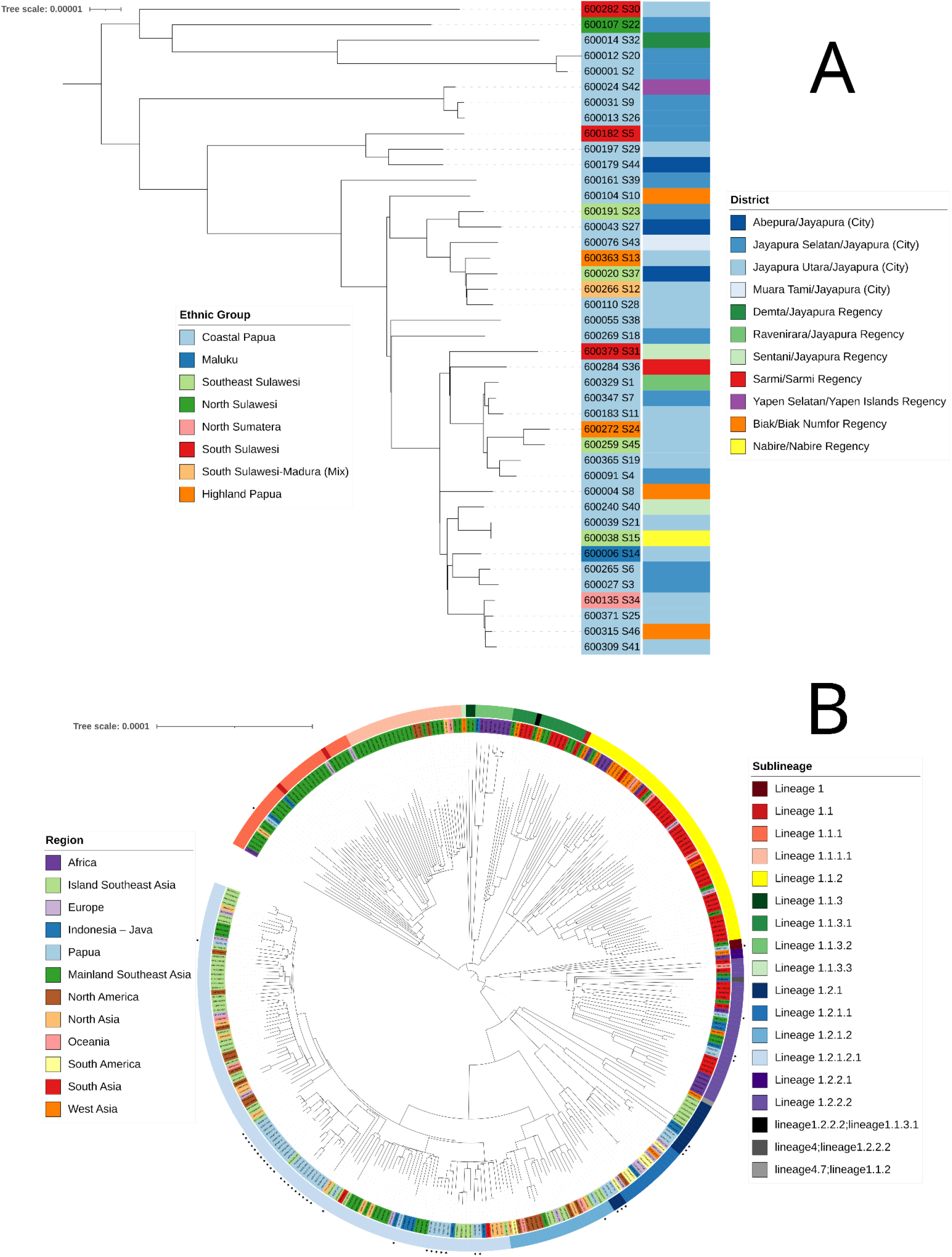
Maximum likelihood phylogenetic tree reconstruction of A) the Papua isolates with the inner color strip indicating patients’ ethnic group and the outer color strip indicating patients’ district of residence and B) the Papua isolates (indicated with dots) with 306 global L1 isolates. The outer ring indicates sublineage information and the inner ring indicates the region of origin.

Only 5 isolates (11.9%) had any mutation associated with drug resistance (**Table 3**). Two had deletions associated with streptomycin resistance. Two had the same substitution in *fabG1* which is associated with isoniazid and ethionamide resistance. One had a substitution in *pncA* resulting in serine substituting glycine, which is associated with pyrazinamide resistance.

**Table 3:** Drug resistance associated mutations from the Papua isolates. Resistance prediction was generated through analysis by TB-Profiler.

| Isolate | Mutation | Associated Drug Resistance |
| --- | --- | --- |
| 600012 | gid_Chromosome:g.4407919_4408026del | Streptomycin |
| 600038 | fabG1_c.-15C>T | Isoniazid - Ethionamide |
| 600039 | fabG1_c.-15C>T | Isoniazid - Ethionamide |
| 600104 | pncA_p.Gly78Ser | Pyrazinamide |
| 600135 | gid_Chromosome:g.4407889_4407959del | Streptomycin |

Based on visual inspection of the phylogenetic tree of the Papuan L1 genomes, there appeared to be no clustering based on geographic location nor ethnicity (**Figure 2**). To investigate this more empirically, we calculated the pairwise SNP distance between each pair of genomes in our sample set. We then compared the distribution of pairwise SNP distances between isolates that were from the same district, and those from different districts, as well as isolate pairs from patients with the same ethnicity and patients from different ethnicities (**Figure 3**). Both density graphs show a bimodal curve, with one broad peak between 0-400 SNPs representing the distances within the L1.2.1.2.1 clade, and another peak between 600-1000 SNPs. There was only one (1/861, 0.1%) closely related pair of isolates (pairs that had a SNP distance less than 10). Twenty-two percent (190/861) of pairs came from patients living in the same district and 48% of pairs came from patients with the same ethnic background (416/861). There was no significant difference in the distances between pairs from the same district compared to those from different districts (Wilcoxon Rank Sum Test, p-value = 0.71) as well as there being no significant difference between pairs from the same ethnicity compared to those from different ethnicities (Wilcoxon Rank Sum Test, p-value = 0.54).

**Figure 3:**
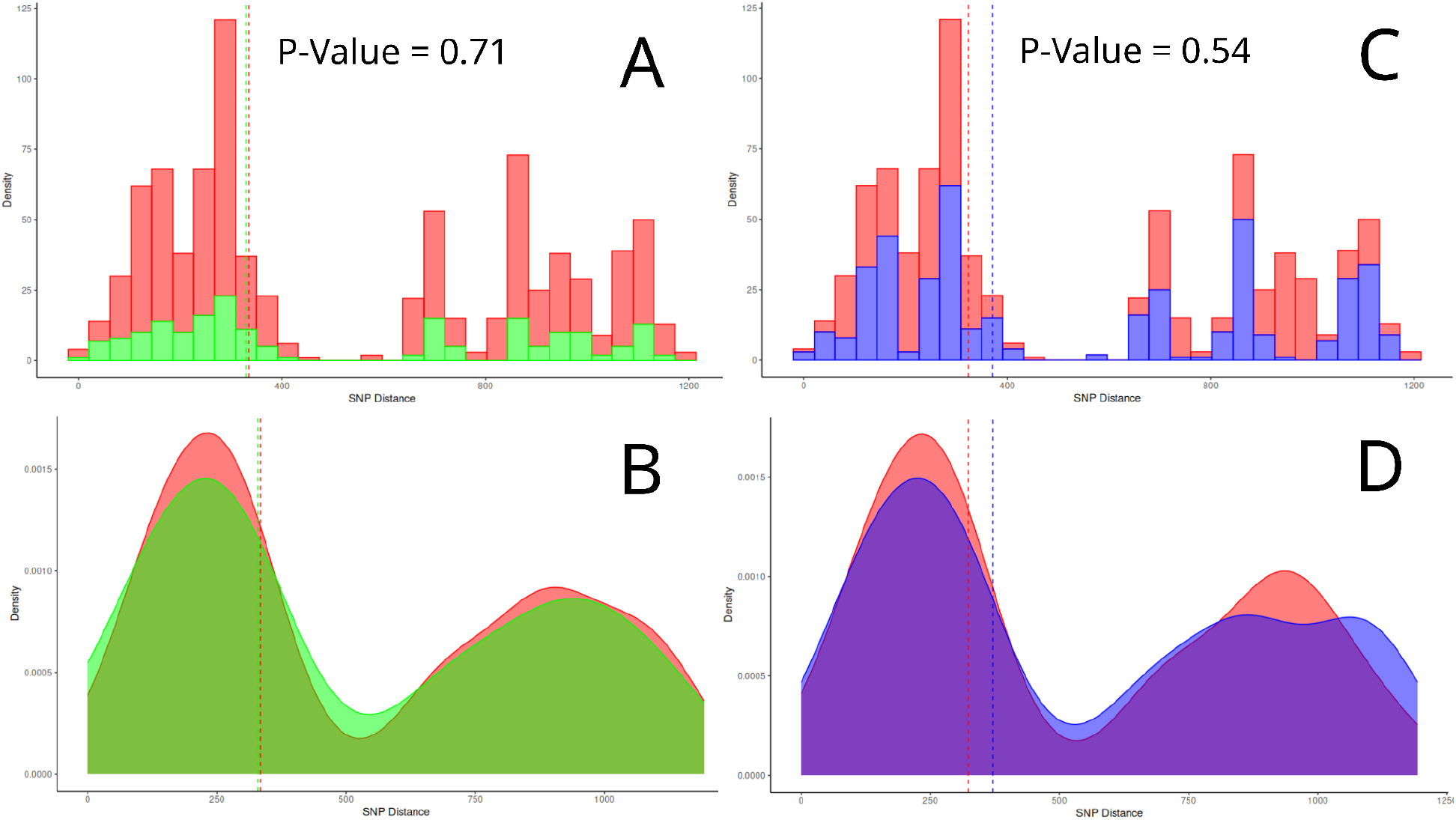
A). Histogram of sample pairs from the same district (Green) and from different districts (Red). B). Density distribution of sample pairs from the same district (Green) and from different districts (Red). C). Histogram of sample pairs from patients with the same ethnic background (Blue) and different ethnic backgrounds (Red). D). Density distribution of sample pairs from patients with the same ethnic background (Blue) and different ethnic backgrounds (Red). Dashed lines indicate median. The Wilcoxon Rank Sum Test was used to compare medians.

When placed into the context of 306 L1 genomes from other studies, the Papuan isolates belong to 13 separate phylogenetic sub-clades, ranging in size between 2-21 (median size = 3). Nine of these sub-clades belong to sublineage L1.2.1 (9/13, 69.2%), three with sublineage L1.2.2 (3/13, 23.1%), and one with sublineage L1.1 (1/13, 7.7%). Of the 13 sub-clades, 5 had isolates from Java (Indonesia) (5/13, 38.5%) as the closest neighbor, 4 were closest with isolates from Island Southeast Asia (4/13, 30.8%), which are Malaysia (2/13, 15.4%), East Timor (1/13, 7.7%), and The Philippines (1/13, 7.7%), 3 were closest with isolates from Mainland Southeast Asia (3/13, 23.1%), which are Cambodia (2/13, 15.4%) and Myanmar (1/13, 7.7%), and 1 was closest with isolates from Sudan in Africa (1/13, 7.7%). The largest phylogenetic group contained 20 Papua isolates along with 1 isolate originating from East Timor (**Figure 2**).

## Discussion

In this study, we provide insights into the genomic characteristics of *Mycobacterium tuberculosis* Lineage 1 from Papua, Indonesia. We found a genetically diverse *MTB* L1 population from patients from across Northern Papua, despite sampling being limited to Jayapura City itself. Despite the wide geographic range and diverse ethnic groups of patients sampled, we were unable to find any genetic clustering by ethnicity or geographic location.

We also observed very few closely related isolates (<10 SNPs pairwise distance), which is consistent with a high burden of disease in Papua, combined with a relatively low sample size. In 2017, there were 3392 reported cases of pulmonary TB in Papua [31.] Assuming that 28% of this is *MTB* L1 [3], we only achieved a sampling fraction of about 0.044. However, considering the limited public access to healthcare in Papua, it is likely that the number of cases is under-reported and the true number is much higher. The lack of closely related pairs indicates that we have not sampled patients from recent transmission chains.

While the small sample size limits our power to detect fine-scale transmission, these data suggest that *MTB* is transmitted between people from different districts and different ethnicities over longer time frames. This is shown by the fact that *MTB* isolates from patients from the same district had a similar pairwise SNP distance compared to patients from different districts. This indicates that there was no evidence that patients are more likely to transmit to people who live in their district compared to people who live outside their district. This is in contrast to Holt, *et al*., [32] who showed that *MTB* L1 isolates from people in the same district were more closely related than *MTB* from people in different districts in Ho Chi Minh City.

The large sub-clade of L1.2.1 isolates from Papua and East Timor in the phylogenetic analysis may show an endemic population, which has been previously speculated to exist in Timor and New Guinea Islands [2]. Our finding that L1.2.1 is common in Papua builds on other research showing that L1.2.1 is common in other Island Southeast Asian Countries. A study in Eastern Malaysia found that of the 208 isolates collected and analyzed, 92% (192/208) belonged to sublineage 1.2.1 [33] and 80% (143/178) of isolates in the Philippines also belonged to this sublineage [34]. However, only 27% (52/192) of the L1.2.1 isolates from Eastern Malaysia belonged to the same L1.2.1.2 sublineage found in Papua, with a majority (122/192, 64%), instead, belonging to sublineage L1.2.1.3 [33]. Although the paper did not specify that the Philippine isolates belonged to sublineage L1.2.1.2 [34], investigation into the SRA database showed a high proportion of L1.2.1.2.1 among *MTB* L1 isolates in the Philippines. This is also shown in the global dataset from the Philippines used in this study.

Thus, it may be implied that most of the isolates from Phelan, *et al*., [34] belong to L1.2.1.2.1 and show a similar population with Papua.

The high proportion of L1 is unique to Eastern Indonesia compared to Western and Central Indonesia where the proportion of L2 and L4 is higher [8–10]. This is also true in neighboring Papua New Guinea, where *MTB* L2 and L4 are the most dominant lineages [35]. However, the situation may be changing, as based on our sampling we observed probable transmissions of *MTB* from the rest of Indonesia to Papua. Therefore, we predict that the proportion of L2 and L4 will increase in Papua [14].

In Mainland Southeast Asia, however, the most dominant *MTB* L1 sublineage is L1.2.1.

Palittapongarnpim, *et al*., [6], Edokimov, *et al*., [29], and Le Hang, *et al*., [36] showed high incidence on L1.1 in Thailand, Cambodia, and Vietnam respectively. While sublineage L1.2.2 can be found across Southeast Asia, the L1.2.2.2 found in the Papua dataset is most commonly found in Thailand [2]. The presence of L1.2.2 and L1.1 in the Papua dataset indicates multiple reintroduction events of *MTB* L1.

The small number of drug-resistant *MTB* could be due to the lack of effective healthcare in the province. Treatment success rate in Papua is between 60-70% and has a high loss to follow up rate (16.4%) [11]. The main reasons cited for non-adherence to TB treatment was the travel cost, the distance to their nearest healthcare facility, and a constantly changing place of residence [11]. Many in the rural areas come to Jayapura for seasonal work or to seek health treatment but many return home before completion of their treatment and are unable to continue treatment there. The lack of effective programs may not provide the necessary selective pressure to force the local *MTB* population to develop drug resistance.

As of writing, we have only managed to find one other study that conducted whole-genome sequencing of *MTB* in Papua [37]. Maladan, *et al*., [37] found a near equal distribution between L1 (7/19, 37%), L2 (5/19. 26%), and L4 (7/19, 37%). However, the study’s sample size is small, only focuses on drug-resistant TB, and lacked information regarding sample sublineage. The small sample size and lack of such studies is not surprising considering Papua’s remote location. Along with the lack of infrastructure and limited access to healthcare, it has all made Papua one of the hardest locations to conduct healthcare research, especially whole-genome sequencing studies on *MTB*.

## Conclusion

In this study, we have focussed on an under-researched lineage in an underdeveloped province of a country with a major burden of *MTB*. We found that the *MTB* L1 population is diverse, has long existed in the region, but has had frequent introductions into the population. However, our sample size is small compared with the burden of disease and only focuses on *MTB* L1. Further studies are needed that include more samples and comparison with other lineages, especially the more widespread L2 and L4.

## Data Availability

All data produced in the present study are available upon reasonable request to the authors

