## Supplemental Tables for "A description of Lineage 1 *Mycobacterium tuberculosis* from Papua, Indonesia"

**Supplementary Materials**

**Table S1:** Region of origin of the *MTB* L1 genomic read sets included in this study

| Location | n | Location | n | Location | n |
| --- | --- | --- | --- | --- | --- |
| ***Southeast Asia*** | ***148*** | ***South Asia*** | ***53*** | ***North Asia*** | ***17*** |
| Indonesia (Java) | 16 | Bangladesh | 10 | China | 1 |
| Papua New Guinea | 6 | India | 20 | Japan | 16 |
| East Timor | 6 | Sri Lanka | 3 | ***West Asia*** | ***18*** |
| Malaysia | 20 | Pakistan | 12 | Iran | 1 |
| Philippines | 20 | Afghanistan | 7 | Palestine | 1 |
| Vietnam | 20 | Nepal | 1 | Yemen | 1 |
| Cambodia | 20 | ***Europe*** | ***16*** | Saudi Arabia | 15 |
| Thailand | 20 | Germany | 8 | ***Africa*** | ***20*** |
| Myanmar | 20 | Netherlands | 4 | Tanzania | 10 |
| ***Oceania*** | ***9*** | Sweden | 3 | Mozambique | 5 |
| Australia | 8 | Switzerland | 1 | Ghana | 1 |
| Kiribati | 1 | ***North America*** | ***20*** | Liberia | 1 |
| ***South America*** | ***7*** | Mexico | 1 | Sudan | 1 |
| Peru | 6 | Canada | 7 | Sierra Leone | 1 |
| Ecuador | 1 | USA | 12 | Uganda | 1 |

**Table S2:** Summary of TB-Profiler results of the Papua and global *MTB* L1 isolates

| Sample | Location | main_lineage | sub_lineage | DR_type |
| --- | --- | --- | --- | --- |
| 600001_S2 | Papua – Jayapura (City) | lineage1 | lineage1.2.2.2 | Sensitive |
| 600004_S8 | Papua – Biak Numfor | lineage1 | lineage1.2.1.2.1 | Sensitive |
| 600006_S14 | Papua – Jayapura (City) | lineage1 | lineage1.2.1.2.1 | Sensitive |
| 600012_S20 | Papua – Jayapura (City) | lineage1 | lineage1.2.2.2 | Other |
| 600013_S26 | Papua – Jayapura (City) | lineage1 | lineage1.2.1 | Sensitive |
| 600014_S32 | Papua – Jayapura (Regency) | lineage1 | lineage1.2.2.2 | Sensitive |
| 600020_S37 | Papua – Jayapura (City) | lineage1 | lineage1.2.1.2.1 | Sensitive |
| 600024_S42 | Papua – Yapen Islands | lineage1 | lineage1.2.1 | Sensitive |
| 600027_S3 | Papua – Jayapura (City) | lineage1 | lineage1.2.1.2.1 | Sensitive |
| 600031_S9 | Papua – Jayapura (City) | lineage1 | lineage1.2.1 | Sensitive |
| 600038_S15 | Papua – Nabire | lineage1 | lineage1.2.1.2.1 | Pre-MDR |
| 600039_S21 | Papua – Jayapura (City) | lineage1 | lineage1.2.1.2.1 | Pre-MDR |
| 600043_S27 | Papua – Jayapura (City) | lineage1 | lineage1.2.1.2.1 | Sensitive |
| 600055_S38 | Papua – Jayapura (City) | lineage1 | lineage1.2.1.2.1 | Sensitive |
| 600076_S43 | Papua – Jayapura (City) | lineage1 | lineage1.2.1.2.1 | Sensitive |
| 600091_S4 | Papua – Jayapura (City) | lineage1 | lineage1.2.1.2.1 | Sensitive |
| 600104_S10 | Papua – Biak Numfor | lineage1 | lineage1.2.1.2.1 | Other |
| 600107_S22 | Papua – Jayapura (City) | lineage1 | lineage1 | Sensitive |
| 600110_S28 | Papua – Jayapura (City) | lineage1 | lineage1.2.1.2.1 | Sensitive |
| 600135_S34 | Papua – Jayapura (City) | lineage1 | lineage1.2.1.2.1 | Other |
| 600161_S39 | Papua – Jayapura (City) | lineage1 | lineage1.2.1.2 | Sensitive |
| 600179_S44 | Papua – Jayapura (City) | lineage1 | lineage1.2.1 | Sensitive |
| 600182_S5 | Papua – Jayapura (City) | lineage1 | lineage1.2.1 | Sensitive |
| 600183_S11 | Papua – Jayapura (City) | lineage1 | lineage1.2.1.2.1 | Sensitive |
| 600191_S23 | Papua – Jayapura (City) | lineage1 | lineage1.2.1.2.1 | Sensitive |
| 600197_S29 | Papua – Jayapura (City) | lineage1 | lineage1.2.1 | Sensitive |
| 600240_S40 | Papua – Jayapura (Regency) | lineage1 | lineage1.2.1.2.1 | Sensitive |
| 600259_S45 | Papua – Jayapura (City) | lineage1 | lineage1.2.1.2.1 | Sensitive |
| 600265_S6 | Papua – Jayapura (City) | lineage1 | lineage1.2.1.2.1 | Sensitive |
| 600266_S12 | Papua – Jayapura (City) | lineage1 | lineage1.2.1.2.1 | Sensitive |
| 600269_S18 | Papua – Jayapura (City) | lineage1 | lineage1.2.1.2.1 | Sensitive |
| 600272_S24 | Papua – Jayapura (City) | lineage1 | lineage1.2.1.2.1 | Sensitive |
| 600282_S30 | Papua – Jayapura (City) | lineage1 | lineage1.1.1 | Sensitive |
| 600284_S36 | Papua – Sarmi | lineage1 | lineage1.2.1.2.1 | Sensitive |
| 600309_S41 | Papua – Jayapura (City) | lineage1 | lineage1.2.1.2.1 | Sensitive |
| 600315_S46 | Papua – Biak Numfor | lineage1 | lineage1.2.1.2.1 | Sensitive |
| 600329_S1 | Papua – Jayapura (Regency) | lineage1 | lineage1.2.1.2.1 | Sensitive |
| 600347_S7 | Papua – Jayapura (City) | lineage1 | lineage1.2.1.2.1 | Sensitive |
| 600363_S13 | Papua – Jayapura (City) | lineage1 | lineage1.2.1.2.1 | Sensitive |
| 600365_S19 | Papua – Jayapura (City) | lineage1 | lineage1.2.1.2.1 | Sensitive |
| 600371_S25 | Papua – Jayapura (City) | lineage1 | lineage1.2.1.2.1 | Sensitive |
| 600379_S31 | Papua – Jayapura (Regency) | lineage1 | lineage1.2.1.2.1 | Sensitive |
| DRR034354 | Japan | lineage1 | lineage1.2.1.2 | Sensitive |
| DRR034365 | Japan | lineage1 | lineage1.2.1.2.1 | Pre-MDR |
| DRR034388 | Japan | lineage1 | lineage1.2.1.2.1 | Sensitive |
| DRR034446 | Japan | lineage1 | lineage1.2.1.2.1 | Sensitive |
| DRR034486 | Japan | lineage1 | lineage1.2.1.2 | Sensitive |
| DRR034522 | Japan | lineage1 | lineage1.2.1.2.1 | Sensitive |
| DRR034523 | Japan | lineage1 | lineage1.2.1.2.1 | Pre-MDR |
| DRR034525 | Japan | lineage1 | lineage1.2.1.2.1 | Pre-MDR |
| DRR034530 | Japan | lineage1 | lineage1.1.1.1 | Sensitive |
| DRR130067 | Japan | lineage1 | lineage1.2.1.2.1 | MDR |
| DRR130068 | Japan | lineage1 | lineage1.2.1.2.1 | MDR |
| DRR130082 | Japan | lineage1 | lineage1.2.1.2 | MDR |
| DRR130094 | Japan | lineage1 | lineage1.2.1.2.1 | MDR |
| DRR130125 | Japan | lineage1 | lineage1.2.1.2.1 | MDR |
| DRR130216 | Japan | lineage1 | lineage1.2.1.2.1 | MDR |
| DRR130222 | Japan | lineage1 | lineage1.2.1.2.1 | Sensitive |
| ERR036236 | Tanzania | lineage1 | lineage1.1.3.2 | Sensitive |
| ERR037496 | Tanzania | lineage1 | lineage1.2.2.1 | Sensitive |
| ERR037511 | Tanzania | lineage1 | lineage1.1.3.2 | Sensitive |
| ERR037519 | Tanzania | lineage1 | lineage1.1.3.2 | Sensitive |
| ERR037520 | Tanzania | lineage1 | lineage1.1.3.2 | Sensitive |
| ERR037555 | Tanzania | lineage1 | lineage1.1.3.2 | Sensitive |
| ERR1023294 | Peru | lineage1 | lineage1.2.1.1 | Pre-MDR |
| ERR1023355 | Peru | lineage1 | lineage1.2.1.1 | Sensitive |
| ERR1023359 | Peru | lineage1 | lineage1.2.1.1 | Sensitive |
| ERR1023370 | Peru | lineage1 | lineage1.2.1.1 | Sensitive |
| ERR1023375 | Peru | lineage1 | lineage1.2.1.1 | Pre-MDR |
| ERR1023388 | Peru | lineage1 | lineage1.2.1.1 | Sensitive |
| ERR1213947 | Bangladesh | lineage1 | lineage1.1.2 | Pre-MDR |
| ERR1213948 | Bangladesh | lineage1 | lineage1.2.2.2 | Pre-MDR |
| ERR1213949 | Bangladesh | lineage1 | lineage1.1.3.1 | Other |
| ERR2199896 | Germany | lineage1 | lineage1.1.2 | Sensitive |
| ERR2200142 | Iran | lineage1 | lineage1.1.2 | Sensitive |
| ERR2200150 | Germany | lineage1 | lineage1.2.1.2.1 | Sensitive |
| ERR234205 | Ghana | lineage1 | lineage1.1.2 | Other |
| ERR245670 | Tanzania | lineage1 | lineage1.2.2.2 | MDR |
| ERR245763 | Tanzania | lineage1 | lineage1.1.3.2 | Sensitive |
| ERR245833 | Tanzania | lineage1 | lineage1.2.2.2 | Sensitive |
| ERR2517163 | Netherlands | lineage1 | lineage1.2.1.1 | Sensitive |
| ERR2517226 | Netherlands | lineage1 | lineage1.2.1.2 | Sensitive |
| ERR2517328 | Netherlands | lineage1 | lineage1.2.1.1 | Other |
| ERR2517538 | Netherlands | lineage1 | lineage1.2.1.1 | Sensitive |
| ERR2660431 | Mozambique | lineage1 | lineage1.2.2.2 | Sensitive |
| ERR2660436 | Mozambique | lineage1 | lineage1.2.2.2 | Sensitive |
| ERR2660437 | Mozambique | lineage1 | lineage1.2.2.2 | Sensitive |
| ERR2660440 | Mozambique | lineage1 | lineage1.1.2 | Pre-MDR |
| ERR2660446 | Mozambique | lineage1 | lineage1.1.3.2 | Sensitive |
| ERR2704679 | China | lineage1 | lineage1.1.1 | Sensitive |
| ERR3077942 | Tanzania | lineage1 | lineage1.1.2 | MDR |
| ERR3148194 | Mexico | lineage1 | lineage1.2.1.2 | MDR |
| ERR3256112 | Philippines | lineage1 | lineage1.2.1.2.1 | MDR |
| ERR3256125 | Philippines | lineage1 | lineage1.2.1.2.1 | Other |
| ERR3256135 | Philippines | lineage1 | lineage1.2.1.2 | MDR |
| ERR3256136 | Philippines | lineage1 | lineage1.2.1.2.1 | Sensitive |
| ERR3256141 | Philippines | lineage1 | lineage1.2.1.2.1 | Pre-MDR |
| ERR3256152 | Philippines | lineage1 | lineage1.2.1.2.1 | Pre-MDR |
| ERR3256155 | Philippines | lineage1 | lineage1.2.1.2.1 | Pre-MDR |
| ERR3256177 | Philippines | lineage1 | lineage1.2.1.2 | MDR |
| ERR3256181 | Philippines | lineage1 | lineage1.2.1.2 | Sensitive |
| ERR3256186 | Philippines | lineage1 | lineage1.2.1.2 | MDR |
| ERR3256188 | Philippines | lineage1 | lineage1.2.1.2.1 | Pre-MDR |
| ERR3256210 | Philippines | lineage1 | lineage1.2.1.2.1 | MDR |
| ERR3256213 | Philippines | lineage1 | lineage1.2.1.2.1 | Sensitive |
| ERR3256214 | Philippines | lineage1 | lineage1.2.1.2.1 | Sensitive |
| ERR3256224 | Philippines | lineage1 | lineage1.2.1.2.1 | Sensitive |
| ERR3256228 | Philippines | lineage1 | lineage1.2.1.2.1 | Sensitive |
| ERR3256249 | Philippines | lineage1 | lineage1.2.1.2.1 | Sensitive |
| ERR3256253 | Philippines | lineage1 | lineage1.2.1.2.1 | Sensitive |
| ERR3256267 | Philippines | lineage1 | lineage1.2.1.2.1 | Pre-MDR |
| ERR3256286 | Philippines | lineage1 | lineage1.2.1.2.1 | Sensitive |
| ERR3324364 | Liberia | lineage1 | lineage1.1.2 | Other |
| ERR3335730 | Pakistan | lineage1 | lineage1.2.2.2 | Other |
| ERR3335746 | Pakistan | lineage1 | lineage1.2.2.2 | Other |
| ERR3335753 | Pakistan | lineage1 | lineage1.1.2 | Pre-MDR |
| ERR3470572 | Sudan | lineage1 | lineage1.2.2.2 | Sensitive |
| ERR4050138 | Germany | lineage1 | lineage1.1.2 | Sensitive |
| ERR550994 | Sierra Leone | lineage1 | lineage1.1.1 | Sensitive |
| ERR551371 | Indonesia | lineage1 | lineage1.2.1.2.1 | Sensitive |
| ERR551435 | Indonesia | lineage1 | lineage4 | lineage4 |
| ERR551865 | Afghanistan | lineage1 | lineage1.1.3.1 | Sensitive |
| ERR551933 | Germany | lineage1 | lineage1.1.1 | Sensitive |
| ERR552076 | Afghanistan | lineage1 | lineage1.1.2 | Sensitive |
| ERR552683 | Germany | lineage1 | lineage1.1.1 | Other |
| ERR552750 | Pakistan | lineage1 | lineage1.1.2 | Sensitive |
| ERR553088 | Indonesia | lineage1 | lineage1.2.1 | Sensitive |
| ERR718265 | Thailand | lineage1 | lineage1.1.3 | Sensitive |
| ERR718367 | Thailand | lineage1 | lineage1.2.1.2.1 | Sensitive |
| ERR718394 | Thailand | lineage1 | lineage1.2.1.2 | Sensitive |
| ERR718458 | Thailand | lineage1 | lineage1.1.3 | Sensitive |
| ERR751660 | Thailand | lineage1 | lineage1.2.1.2.1 | Sensitive |
| ERR751690 | Thailand | lineage1 | lineage1.1.1 | Other |
| ERR751698 | Thailand | lineage1 | lineage1.1.1.1 | Sensitive |
| ERR751739 | Thailand | lineage1 | lineage1.2.2.2 | Sensitive |
| ERR751746 | Thailand | lineage1 | lineage1.2.1.2.1 | Sensitive |
| ERR751797 | Thailand | lineage1 | lineage1.2.1.2.1 | Sensitive |
| ERR751841 | Thailand | lineage1 | lineage1.1.1 | Pre-MDR |
| ERR752059 | Thailand | lineage1 | lineage1.2.1.2.1 | Sensitive |
| ERR752133 | Thailand | lineage1 | lineage1.1.1 | Sensitive |
| ERR752187 | Thailand | lineage1 | lineage1.1.1 | Sensitive |
| ERR752209 | Thailand | lineage1 | lineage1.1.1 | Sensitive |
| ERR767997 | Thailand | lineage1 | lineage1.1.1 | Pre-MDR |
| ERR768010 | Thailand | lineage1 | lineage1.1.1 | Sensitive |
| ERR768016 | Thailand | lineage1 | lineage1.1.1 | Sensitive |
| ERR768038 | Thailand | lineage1 | lineage1.1 | Sensitive |
| ERR7902589 | Pakistan | lineage1 | lineage1.1.2 | Pre-MDR |
| ERR7902598 | Germany | lineage1 | lineage1.2.1.2.1 | Sensitive |
| ERR7902602 | Pakistan | lineage1 | lineage1.2.2.2 | Sensitive |
| ERR7902603 | Bangladesh | lineage1 | lineage1.2.2.2 | Sensitive |
| ERR7902605 | Bangladesh | lineage1 | lineage1.1.3.1 | Sensitive |
| ERR7902613 | Pakistan | lineage1 | lineage1.1.2 | Sensitive |
| ERR7902629 | Afghanistan | lineage1 | lineage1.1.3.1 | Sensitive |
| ERR7902633 | Germany | lineage1 | lineage1.2.1.2.1 | Sensitive |
| ERR7902635 | Pakistan | lineage1 | lineage1.1.2 | Sensitive |
| ERR7902637 | Bangladesh | lineage1 | lineage1.1.3.1 | Sensitive |
| ERR7902638 | Pakistan | lineage1 | lineage1.1.2 | Sensitive |
| ERR7902640 | Saudi Arabia | lineage1 | lineage1.1.3.1 | Other |
| ERR7902641 | Afghanistan | lineage1 | lineage1.1.2 | Sensitive |
| ERR7902652 | Palestine | lineage1 | lineage1.1.3.3 | Sensitive |
| ERR7902661 | Bangladesh | lineage1 | lineage1.1.3.1 | Sensitive |
| ERR7902674 | Germany | lineage1 | lineage1.2.1.2.1 | Other |
| ERR7902689 | Pakistan | lineage1 | lineage1.2.2.2 | Sensitive |
| ERR7902690 | Sri Lanka | lineage1 | lineage1.2.2.2 | Sensitive |
| ERR7902692 | Pakistan | lineage1 | lineage1.2.1.2.1 | Sensitive |
| ERR7902703 | Bangladesh | lineage1 | lineage1.1.2 | Sensitive |
| ERR7902705 | Afghanistan | lineage1 | lineage1.2.2.2 | Sensitive |
| ERR7902706 | Myanmar | lineage1 | lineage1.1.2 | Sensitive |
| ERR7902715 | Kiribati | lineage1 | lineage1.2.1.2 | Sensitive |
| ERR7902723 | Sri Lanka | lineage1 | lineage1.1.2 | Sensitive |
| ERR7902734 | Afghanistan | lineage1 | lineage1.2.2.2 | Sensitive |
| ERR7902738 | Pakistan | lineage1 | lineage1.2.2.2 | Sensitive |
| ERR7902742 | Bangladesh | lineage1 | lineage1.1.3.1 | Sensitive |
| ERR7902743 | Sri Lanka | lineage1 | lineage1.1.2 | Sensitive |
| ERR7902749 | Afghanistan | lineage1 | lineage1.1.2 | Sensitive |
| ERR7902750 | Saudi Arabia | lineage1 | lineage1.1.2 | MDR |
| ERR7902751 | Saudi Arabia | lineage1 | lineage1.2.2.2 | Pre-MDR |
| ERR7902753 | Saudi Arabia | lineage1 | lineage1.1.2 | Other |
| ERR7902754 | Bangladesh | lineage1 | lineage1.1.3.1 | Sensitive |
| ERR7902756 | Indonesia | lineage1 | lineage1.1.3.2 | Sensitive |
| ERR7902757 | Saudi Arabia | lineage1 | lineage1.1.3.1 | Pre-MDR |
| ERR7902758 | Saudi Arabia | lineage1 | lineage1.1.2 | Pre-MDR |
| ERR7902759 | Indonesia | lineage1 | lineage1.2.1 | Sensitive |
| ERR7902760 | Nepal | lineage1 | lineage1.1.2 | Sensitive |
| ERR7902761 | Saudi Arabia | lineage1 | lineage1.1.2 | Sensitive |
| ERR7902762 | Indonesia | lineage1 | lineage1.2.1.1 | Sensitive |
| ERR7902763 | Saudi Arabia | lineage1 | lineage4 | Sensitive |
| ERR7902765 | Saudi Arabia | lineage1 | lineage1.1.2 | Sensitive |
| ERR7902766 | Yemen | lineage1 | lineage1.2.2.1 | Other |
| ERR7902767 | Saudi Arabia | lineage1 | lineage1.2.1.1 | MDR |
| ERR7902768 | Saudi Arabia | lineage1 | lineage1.1.2 | Sensitive |
| ERR7902769 | Saudi Arabia | lineage1 | lineage1.1.2 | MDR |
| ERR7902770 | Saudi Arabia | lineage1 | lineage1.1.2 | MDR |
| ERR7902771 | Indonesia | lineage1 | lineage1.2.1.2.1 | Pre-MDR |
| ERR846991 | Thailand | lineage1 | lineage1.2.1.2.1 | Sensitive |
| ERR983237 | Myanmar | lineage1 | lineage1.1.2 | MDR |
| SRR10214505 | Malaysia | lineage1 | lineage1.2.1.2 | Sensitive |
| SRR10214506 | Malaysia | lineage1 | lineage1.2.1.2.1 | Sensitive |
| SRR10214508 | Malaysia | lineage1 | lineage1.2.1.2.1 | Sensitive |
| SRR1062894 | Uganda | lineage1 | lineage1.1.2 | MDR |
| SRR1144786 | Sweden | lineage1 | lineage1.2.1.1 | Sensitive |
| SRR1165515 | Sweden | lineage1 | lineage1.2.1.1 | MDR |
| SRR1165585 | Sweden | lineage1 | lineage1.2.1.1 | MDR |
| SRR1169482 | India | lineage1 | lineage1.1.2 | Sensitive |
| SRR1169548 | India | lineage1 | lineage1.1.2 | Sensitive |
| SRR1169554 | India | lineage1 | lineage1.1.2 | Sensitive |
| SRR1169564 | India | lineage1 | lineage1.1.2 | Sensitive |
| SRR1169570 | India | lineage1 | lineage1.1.2 | Pre-MDR |
| SRR1172716 | India | lineage1 | lineage1.1.2 | Pre-MDR |
| SRR1172787 | India | lineage1 | lineage1.2.2.2 | Sensitive |
| SRR1172799 | India | lineage1 | lineage1.1.2 | Sensitive |
| SRR1172860 | India | lineage1 | lineage1.1.2 | Sensitive |
| SRR1172993 | India | lineage1 | lineage1.1.2 | Sensitive |
| SRR1173013 | India | lineage1 | lineage1.1.2 | Sensitive |
| SRR1173127 | India | lineage1 | lineage1.1.2 | Sensitive |
| SRR1173257 | India | lineage1 | lineage1.1.2 | MDR |
| SRR12006519 | Malaysia | lineage1 | lineage1.2.1.2.1 | Sensitive |
| SRR12006540 | Malaysia | lineage1 | lineage1.2.1.2.1 | Sensitive |
| SRR12006568 | Malaysia | lineage1 | lineage1.2.1.2.1 | Sensitive |
| SRR12006573 | Malaysia | lineage1 | lineage1.2.1.2.1 | Sensitive |
| SRR12006581 | Malaysia | lineage1 | lineage1.2.1.2.1 | Sensitive |
| SRR12006592 | Malaysia | lineage1 | lineage1.2.1.2.1 | Other |
| SRR12006595 | Malaysia | lineage1 | lineage1.2.1.2.1 | Sensitive |
| SRR12006600 | Malaysia | lineage1 | lineage1.2.1.2 | Sensitive |
| SRR12006611 | Malaysia | lineage1 | lineage1.2.1.2.1 | Pre-MDR |
| SRR12006639 | Malaysia | lineage1 | lineage1.2.1.2.1 | Sensitive |
| SRR12006646 | Malaysia | lineage1 | lineage1.2.1.2 | Sensitive |
| SRR12006648 | Malaysia | lineage1 | lineage1.2.1.2.1 | Sensitive |
| SRR12006657 | Malaysia | lineage1 | lineage1.2.1.2.1 | Sensitive |
| SRR12006658 | Malaysia | lineage1 | lineage1.2.1.2.1 | Sensitive |
| SRR12006683 | Malaysia | lineage1 | lineage1.2.1.2.1 | Sensitive |
| SRR12006684 | Malaysia | lineage1 | lineage1.2.1.2.1 | Sensitive |
| SRR12006692 | Malaysia | lineage1 | lineage1.2.1.2.1 | Sensitive |
| SRR12199383 | Myanmar | lineage1 | lineage1.1.2 | Other |
| SRR12199388 | Myanmar | lineage1 | lineage1.2.1.2.1 | Sensitive |
| SRR12199391 | Myanmar | lineage1 | lineage1.1.1 | Sensitive |
| SRR12199395 | Myanmar | lineage1 | lineage1.2.2.2 | Sensitive |
| SRR12199402 | Myanmar | lineage1 | lineage1.1.3.1 | Sensitive |
| SRR12199409 | Myanmar | lineage1 | lineage1.2.2.2 | Other |
| SRR12199411 | Myanmar | lineage1 | lineage1.1.3.1 | Sensitive |
| SRR12199412 | Myanmar | lineage1 | lineage1.2.2.2 | Sensitive |
| SRR12199423 | Myanmar | lineage1 | lineage1.2.2.2 | Sensitive |
| SRR12199437 | Myanmar | lineage1 | lineage1.1.2 | Sensitive |
| SRR12199452 | Myanmar | lineage1 | lineage1.2.1.2.1 | Sensitive |
| SRR12199453 | Myanmar | lineage1 | lineage1 | Other |
| SRR12199456 | Myanmar | lineage1 | lineage1.1 | Sensitive |
| SRR12199458 | Myanmar | lineage1 | lineage1.1.3.1 | Sensitive |
| SRR12199459 | Myanmar | lineage1 | lineage1.1 | Sensitive |
| SRR12199471 | Myanmar | lineage1 | lineage1.1.3.1 | Sensitive |
| SRR12199474 | Myanmar | lineage1 | lineage1.1.3.1 | Sensitive |
| SRR12199479 | Myanmar | lineage1 | lineage1.2.2.2 | Sensitive |
| SRR12882030 | East Timor | lineage1 | lineage1.2.1 | Sensitive |
| SRR12882038 | Indonesia | lineage1 | lineage1.2.1.2.1 | Other |
| SRR12882045 | East Timor | lineage1 | lineage1.2.1 | Sensitive |
| SRR12882047 | East Timor | lineage1 | lineage1.2.1 | Sensitive |
| SRR12882048 | East Timor | lineage1 | lineage1.2.1 | Sensitive |
| SRR12882049 | PNG | lineage1 | lineage1.2.1.2.1 | Sensitive |
| SRR12882260 | PNG | lineage1 | lineage1.2.1 | Sensitive |
| SRR12882272 | PNG | lineage1 | lineage1.2.1 | Sensitive |
| SRR12882278 | PNG | lineage1 | lineage1.2.1.2.1 | Sensitive |
| SRR12882282 | East Timor | lineage1 | lineage1.2.1.2.1 | Sensitive |
| SRR12882292 | Switzerland | lineage1 | lineage1.2.1.2.1 | Sensitive |
| SRR12882483 | Ecuador | lineage1 | lineage1.2.1.2 | Sensitive |
| SRR12882752 | PNG | lineage1 | lineage1.2.1.2.1 | Sensitive |
| SRR12882763 | East Timor | lineage1 | lineage1.2.1 | Sensitive |
| SRR12882771 | PNG | lineage1 | lineage1.2.1.2.1 | Sensitive |
| SRR17729341 | Cambodia | lineage1 | lineage1.1.1 | Other |
| SRR17729344 | Cambodia | lineage1 | lineage1.2.1.2.1 | Sensitive |
| SRR17729345 | Cambodia | lineage1 | lineage1.1.1 | Sensitive |
| SRR17729346 | Cambodia | lineage1 | lineage1.1.1 | Other |
| SRR17729350 | Cambodia | lineage1 | lineage1.1.1 | Sensitive |
| SRR17729353 | Cambodia | lineage1 | lineage1.1.1 | Sensitive |
| SRR17729357 | Cambodia | lineage1 | lineage1.1.1 | Sensitive |
| SRR17729359 | Cambodia | lineage1 | lineage1.2.1.2.1 | Sensitive |
| SRR17729361 | Cambodia | lineage1 | lineage1.2.1.2.1 | Sensitive |
| SRR17729362 | Cambodia | lineage1 | lineage1.1.1 | Sensitive |
| SRR17729375 | Cambodia | lineage1 | lineage1.2.1.2.1 | Sensitive |
| SRR17729376 | Cambodia | lineage1 | lineage1.1.1 | Sensitive |
| SRR17729382 | Cambodia | lineage1 | lineage1.1.1 | Other |
| SRR17729386 | Cambodia | lineage1 | lineage1.1.1 | Sensitive |
| SRR17729396 | Cambodia | lineage1 | lineage1.1.1 | Sensitive |
| SRR17729397 | Cambodia | lineage1 | lineage1.1.1 | Sensitive |
| SRR17729398 | Cambodia | lineage1 | lineage1.2.1.2.1 | Sensitive |
| SRR17729404 | Cambodia | lineage1 | lineage1.2.1.2.1 | Sensitive |
| SRR17729413 | Cambodia | lineage1 | lineage1.1.1 | Sensitive |
| SRR17729419 | Cambodia | lineage1 | lineage1.1.1 | Sensitive |
| SRR5065201 | Vietnam | lineage1 | lineage1.1.1.1 | Sensitive |
| SRR5065243 | Vietnam | lineage1 | lineage1.1.1.1 | Sensitive |
| SRR5065280 | Vietnam | lineage1 | lineage1.1.1.1 | Other |
| SRR5065289 | Vietnam | lineage1 | lineage1.1.1.1 | Pre-MDR |
| SRR5065309 | Vietnam | lineage1 | lineage1.1.1.1 | Sensitive |
| SRR5065386 | Vietnam | lineage1 | lineage1.1.1 | Other |
| SRR5065428 | Vietnam | lineage1 | lineage1.1.1.1 | Sensitive |
| SRR5065491 | Vietnam | lineage1 | lineage1.1.1.1 | Sensitive |
| SRR5065523 | Vietnam | lineage1 | lineage1.1.1.1 | Sensitive |
| SRR5065603 | Vietnam | lineage1 | lineage1.1.1.1 | Sensitive |
| SRR5067252 | Vietnam | lineage1 | lineage1.1.1.1 | Sensitive |
| SRR5067283 | Vietnam | lineage1 | lineage1.1.1.1 | Sensitive |
| SRR5067397 | Vietnam | lineage1 | lineage1.1.1.1 | Sensitive |
| SRR5067445 | Vietnam | lineage1 | lineage1.1.1.1 | Sensitive |
| SRR5067522 | Vietnam | lineage1 | lineage1.1.1.1 | Sensitive |
| SRR5067645 | Vietnam | lineage1 | lineage1.1.1.1 | Pre-MDR |
| SRR5073587 | Vietnam | lineage1 | lineage1.1.1.1 | Sensitive |
| SRR5074086 | Vietnam | lineage1 | lineage1.1.1.1 | Sensitive |
| SRR5074099 | Vietnam | lineage1 | lineage1.1.1.1 | Sensitive |
| SRR5074124 | Vietnam | lineage1 | lineage1.1.1.1 | Sensitive |
| SRR5341272 | India | lineage1 | lineage1.2.1.2.1 | Pre-MDR |
| SRR5817476 | Australia | lineage1 | lineage1.1.2 | XDR |
| SRR5817479 | Australia | lineage1 | lineage1.2.1.2 | MDR |
| SRR5817481 | Australia | lineage1 | lineage1.2.1.2.1 | Pre-MDR |
| SRR6152694 | Canada | lineage1 | lineage1.2.1.2 | Sensitive |
| SRR6152796 | Canada | lineage1 | lineage1.2.1.2 | Sensitive |
| SRR6152810 | Canada | lineage1 | lineage1.2.1.2 | Sensitive |
| SRR6152816 | Canada | lineage1 | lineage1.2.1.2 | Sensitive |
| SRR6153258 | Canada | lineage1 | lineage1.2.1.2 | Pre-MDR |
| SRR6339636 | Australia | lineage1 | lineage1.2.1.2.1 | MDR |
| SRR6339657 | Australia | lineage1 | lineage1.1.2 | Sensitive |
| SRR6367396 | Australia | lineage1 | lineage1.2.2.2 | Sensitive |
| SRR6367399 | Australia | lineage1 | lineage1.1.2 | MDR |
| SRR6369876 | Australia | lineage1 | lineage1.1.1.1 | MDR |
| SRR6397921 | Canada | lineage1 | lineage1.2.1.1 | Sensitive |
| SRR6397949 | Canada | lineage1 | lineage1.2.1.2 | MDR |
| SRR6480334 | Indonesia – Java | lineage1 | lineage1.2.1.2.1 | Sensitive |
| SRR6480337 | Indonesia – Java | lineage1 | lineage1.2.2.2 | Sensitive |
| SRR6480388 | Indonesia – Java | lineage1 | lineage1.1.1 | Sensitive |
| SRR6480442 | Indonesia – Java | lineage1 | lineage1.2.1.2.1 | Other |
| SRR6480513 | Indonesia – Java | lineage1 | lineage1.2.1.2.1 | Sensitive |
| SRR6480514 | Indonesia – Java | lineage1 | lineage1.2.2.2 | Sensitive |
| SRR6480525 | Indonesia – Java | lineage1 | lineage1.2.2.2 | Sensitive |
| SRR6480622 | Indonesia – Java | lineage1 | lineage1.1.1 | Sensitive |
| SRR7496480 | USA | lineage1 | lineage1.2.1.2.1 | Sensitive |
| SRR7496485 | USA | lineage1 | lineage1.1.1.1 | Pre-MDR |
| SRR7496486 | USA | lineage1 | lineage1.1.1.1 | Pre-MDR |
| SRR7496490 | USA | lineage1 | lineage1.2.1.2.1 | Sensitive |
| SRR7496504 | USA | lineage1 | lineage1.2.1.2.1 | Sensitive |
| SRR7496510 | USA | lineage1 | lineage1.2.1.2.1 | Sensitive |
| SRR7496520 | USA | lineage1 | lineage1.2.1.2.1 | Sensitive |
| SRR7496526 | USA | lineage1 | lineage1.1.1.1 | Sensitive |
| SRR7496532 | USA | lineage1 | lineage1.2.1.2.1 | Sensitive |
| SRR7496534 | USA | lineage1 | lineage1.2.1.2.1 | Sensitive |
| SRR7496547 | USA | lineage1 | lineage1.2.1.2.1 | Sensitive |
| SRR7496548 | USA | lineage1 | lineage1.2.1.2.1 | Sensitive |
| SRR8380861 | India | lineage1 | lineage1.1.2 | Sensitive |
| SRR8380885 | India | lineage1 | lineage1.1.2 | Sensitive |
| SRR8380886 | India | lineage1 | lineage1.1.2 | Sensitive |
| SRR8380889 | India | lineage1 | lineage1.1.2 | Sensitive |
| SRR8380909 | India | lineage1 | lineage1.1.2 | Pre-MDR |
| SRR8380918 | India | lineage1 | lineage1.1.2 | Sensitive |
